# Google Searches for Taste and Smell Loss Anticipate Covid-19 Epidemiology

**DOI:** 10.1101/2020.11.09.20228510

**Authors:** Giuseppe Lippi, Brandon M. Henry, Camilla Mattiuzzi, Fabian Sanchis-Gomar

## Abstract

**Background:** As evidence emerged that loss of taste and/or loss of smell is frequently triggered by severe acute respiratory syndrome coronavirus 2 (SARS-CoV-2) infection, we investigated whether Google searches volume for these two disease-specific symptoms could be associated with disease epidemiology in United States (US).

**Materials and Methods:** We performed an electronic search in Google Trends using the keywords “taste loss” and “smell loss” within the US. The Google searches volume was correlated with the number of new weekly cases of coronavirus disease 2019 (COVID-19) in the country.

**Results:** The weekly Google searches for taste and smell loss exhibited a trend similar to that of new weekly SARS-CoV-2 infections in the US. A nearly perfect correlation was found between Google Trends scores of taste and smell loss (r=0.98; 95% CI, 0.97-0.99; p<0.001). Although a significant association was found between Google searches for the two symptoms and the concomitant number of new weekly SARS-CoV-2 infections reported during the same week, the correlation improved over time. The highest correlation was found comparing Google Trends scores for taste or smell loss and the number of new weekly SARS-CoV-2 infections two weeks later. The correlation coefficient of summing Google Trends scores for the two symptoms and the number of new weekly SARS-CoV-2 infections two weeks later was 0.82 (95% CI, 0.68-0.90; p<0.001), and was associated 0.89 diagnostic accuracy.

**Conclusions:** These findings suggest that Google searches numbers for olfactory and gustatory dysfunctions may help predicting the epidemiological trajectory of COVID-19 early before official reporting.

## Introduction

The Coronavirus Disease 2019 (COVID-19) pandemic outbreak can now be regarded as one of the largest calamities to impact humanity in modern times [1]. Responsible for over 1.3 million deaths to date and following an epidemic trend that is predicted to become even worse in the near future, severe acute respiratory distress syndrome coronavirus 2 (SARS-CoV-2) has already disrupted health care organizations, economies and societies worldwide [2]. With natural herd immunity universally considered unfeasible or virtually unreachable without a concomitant collapse of health care systems and an immense number of further casualties [3], most of our hopes rely on the development of efficient vaccines. Nonetheless, this strategy will expectedly encounter unprecedented challenges due to the need to generate and dispense an efficient and durable immunization to billions of people worldwide [4]. It is predictable that we will need to learn to live with this novel coronavirus for quite a while, adapting our lifestyle and attempting to predict, or at least rapidly identify, manage, and contain new outbreaks or local resurgence [5].

According to the US Centers for Disease Control and Prevention (CDC), the most frequent symptoms reported by patients with SARS-CoV-2 infection are fever, cough, shortness of breath, myalgia, and headache [6]. Nonetheless, evidence has accumulated showing that two atypical symptoms, the loss of taste and/or loss of smell, may be frequently triggered by SARS-CoV-2 infection [7]. This is supported by some recent meta-analyses, such as those published by Hajikhani et al. [8], as well as by Tong and colleagues [9], who reported that olfactory and gustatory dysfunctions are present in over 50% of patients with SARS-CoV-2 infection. Notably, the recent meta-analysis of Hoang et al. also revealed that olfactory and/or gustatory dysfunctions might also be highly specific symptoms of SARS-CoV-2 infection, since they were found to be 10-fold more likely to occur in COVID-19 patients as opposed to those with other acute respiratory infections (odds ratio, 11.3; 95% confidence interval [95% CI], 5.4-23.4) [10]. This would hence suggest that the onset of taste (hypogeusia or ageusia) or smell (i.e., hyposmia or anosmia) dysfunctions should raise high clinical suspicion for the potential of active SARS-CoV-2 infection, thus accelerating diagnosis, isolation and treatment (when necessary) of such patients. Another critical aspect directly related to the appearance of these symptoms is that they may present a unique opportunity for monitoring disease epidemiology that can help predict, anticipate or identify SARS-CoV-2 outbreaks or resurgence within the community, even before specific diagnostic testing is initiated. This would be extremely important in low resource settings, where widespread molecular testing is impossible or unfeasible, as well as in the northern hemisphere, as it heads into the winter months, concomitant with peak cold and flu season.

## Materials and Methods

We performed an electronic search in Google Trends (Google Inc. Mountain View, CA, US), using the keywords “taste loss” AND “smell loss,” limited with country option set to “United States.” These search terms were selected as they represent highly paradigmatic symptoms reported at disease onset in patients with SARS-CoV-2 infection [7]. The search period ranged between the third week of January 2020 and present time (i.e., between January 20, 2020 and November 1, 2020). The weekly Google Trends score, reflecting the mean volume of Google searches for a specific term in the preceding seven days, was downloaded and imported into a Microsoft Excel file (Microsoft, Redmond, WA, United States). The number of new weekly cases of COVID-19 in the US was retrieved from the official website of the CDC [11], and tabulated into the same Excel worksheet. The Google search volume for the two keywords and the new weekly SARS-CoV-2 infections in the US were analyzed with Spearman’s correlation, whilst the diagnostic performance was assessed by receiver operating characteristics (ROC) curves. The statistical analysis was carried out using Analyse-it (Analyse-it Software Ltd, Leeds, UK). The study was conducted in accordance with the Declaration of Helsinki, under the terms of relevant local legislation.

## Results

A nearly perfect correlation was found between the Google Trends scores of taste loss and smell loss (r=0.98; 95% CI, 0.97-0.99; p<0.001) (Figure 1). The trends of weekly Google searches for taste and smell loss and the number of new weekly SARS-CoV-2 infections cases in the US are shown in Figure 2. The three curves exhibited a relatively similar shape. However, changes in Google search trends for taste and smell loss appeared to occur slightly earlier in comparison to changes in the number of weekly SARS-CoV-2 infections. The Spearman’s correlations between Google Trends score for taste or smell loss and the number of new weekly SARS-CoV-2 infections in the US reported during the same period of the Google searches (same week, i.e. index week), as well as the correlations between Google Trend scores in the index week versus the number of cases reported in the subsequent 1, 2, 3 and 4 weeks after are shown in Table 1. Although a significant association was found between the volume of Google searches for the two keywords and the number of concomitant new weekly SARS-CoV-2 infections diagnosed during the same week, the correlation gradually improved over time. Google Trends scores for taste loss or smell loss from the index week were correlated with the number of new weekly SARS-CoV-2 infections in the following four weeks. The highest correlation was found comparing Google Trends scores for taste loss or smell loss at the index week and the number of new weekly SARS-CoV-2 infections reported in the US two weeks later (Table 1). An even stronger correlation was found when the sum of the Google Trends scores for taste loss or smell loss at the index week was compared with newly diagnosed weekly SARS-CoV-2 infections two weeks afterwards (r=0.82; 95% CI, 0.68-0.90; p<0.001) (Figure 3). A positive variation (i.e., an increase from the previous week) in the number of Google searches for taste or smell loss was characterized by an area under the curve (AUC) of 0.89 (95% CI, 0.79-0.99; 0.83 sensitivity and 0.70 specificity) for predicting an increasing trend of new SARS-CoV-2 weekly infections two weeks after the date of Google searches.

**Table 1.**
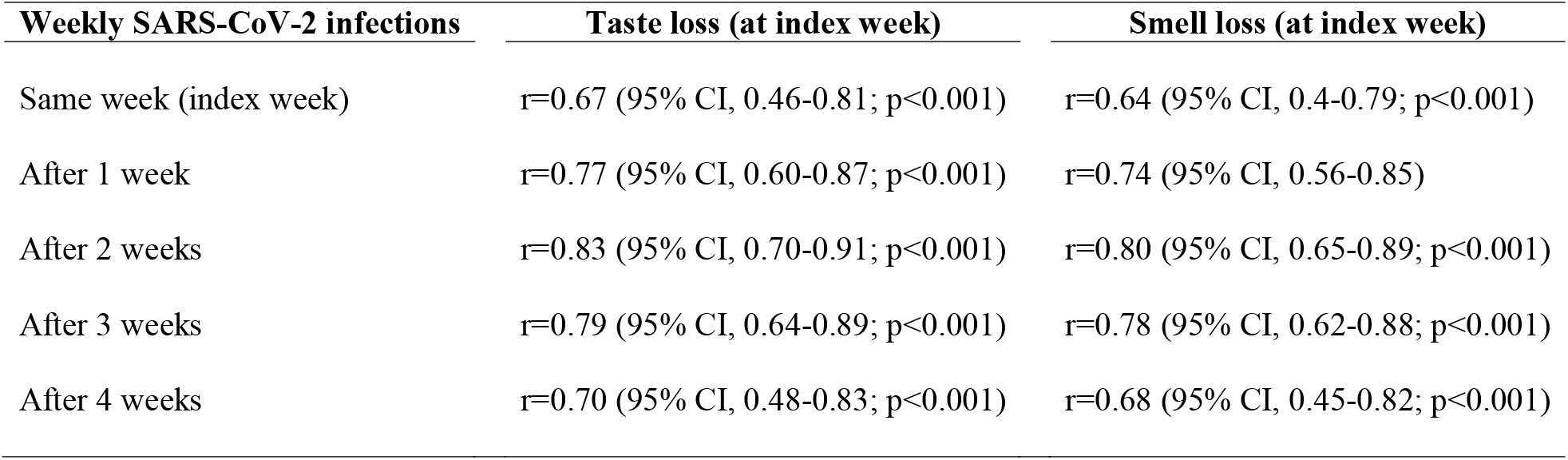
Spearman’s correlation between Google Trends scores for the keywords “taste loss” or “smell loss” and the number of new weekly SARS-CoV-2 infections in the US reported during the same period of the Google searches (index week), as well as the Google Trend scores from the index week and the number of new weekly SARS-CoV-2 cases in the USA in the following 1, 2, 3 and 4 weeks. Data are between January 20, 2020 and November 1, 2020.

**Figure 1.**
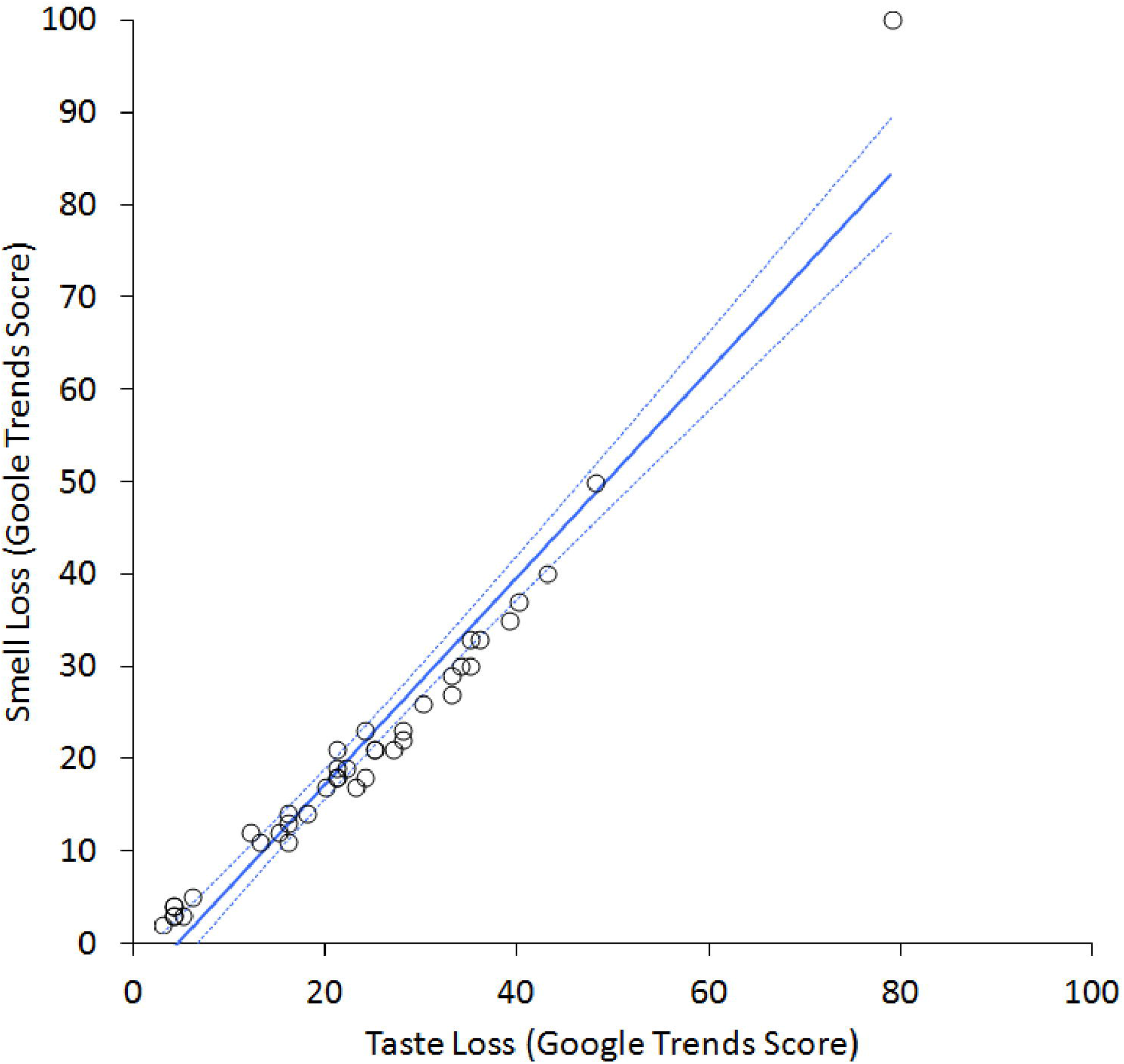
Spearman’s correlation between Google search volumes for “loss of taste” and “loss of smell” in the US, between January 20, 2020 and November 1, 2020.

**Figure 2.**
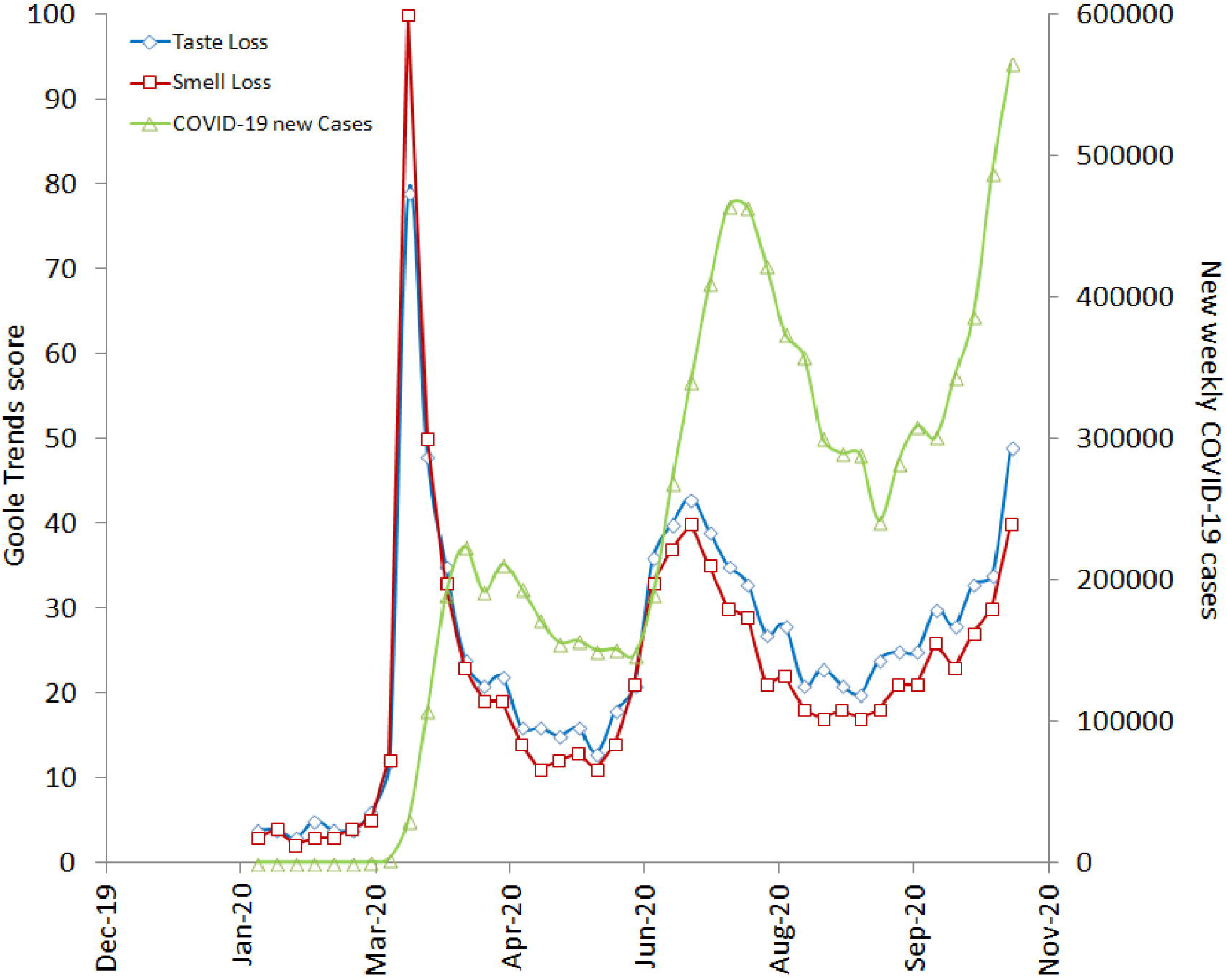
Number of newly diagnosed SARS-CoV-2 infections per week and weekly volume of Google searches (Google Trends score) for “loss of taste”, and “loss of smell” in the US, between January 20, 2020 and November 1, 2020.

**Figure 3.**
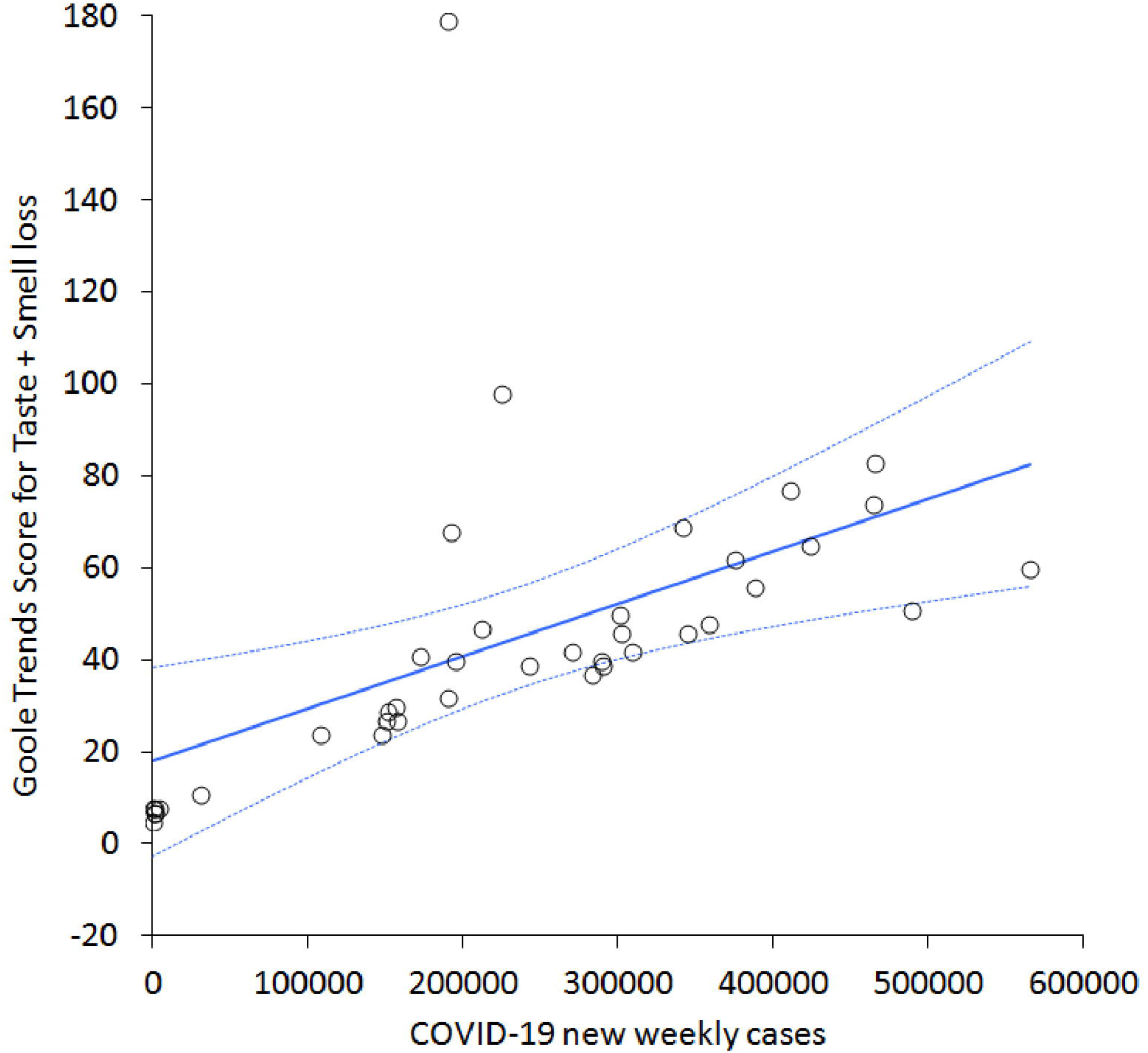
Spearman’s correlation between the sum of Google searches for “loss of taste” and “loss of smell” and the number of new SARS-CoV-2 weekly cases two weeks later in the US, between January 20, 2020 and November 1, 2020.

## Discussion

Evidence has demonstrated that when frequent and specific symptoms characterize a particular pathology, including infectious diseases, real-time analysis of web searches may help anticipate or identify the epidemiological trajectory of that disease within a specific geographic settings [12]. With respect to respiratory diseases, it has been earlier shown that the volume of Google searches for influenza may reliably mirror the real-time evolution of disease at many temporal and spatial resolutions [13,14]. Similar evidence has been reported for other common respiratory infections, such as those caused by Respiratory Syncytial Virus [15] and even for the previous coronavirus disease Middle East Acute Respiratory Syndrome (MERS) [16]. This new branch of epidemiology, which has been informally called “infodemiology,” is defined as a science based on using electronic Web-based information to inform and guide public health policies [17] and is increasingly used to support conventional epidemiology in preventing, containing and managing COVID-19 around the world [18,19].

Interestingly, Jimenez et al. recently showed that Google Trends search data for a combination of symptoms such as fatigue, diarrhea, sore throat, fever, pneumonia, lost sense of smell, and cough could efficiently predict the evolution of the COVID-19 outbreak in Spain with 11 days of anticipation [20]. Similar reports have been published in other countries, though most of these have included a vast array of symptoms, such as fever, cough, and dyspnea, which are not specific for COVID-19 [21,22]. However, as winter arrives in northern hemisphere bringing with it cold and flu season, these non-specific symptoms may be of limited utility as search terms for epidemiologic monitoring. Conversely, the onset of olfactory and gustatory dysfunctions, especially in combination, is rare in patients with other upper respiratory viral infections [23]. The presence of these symptoms presents a high likelihood of being linked to an ongoing or recent SARS-CoV-2 infection [24], which likely encourages patients to undergo molecular testing and initiate social distancing and isolation [25]. Multiple potential cellular and molecular causes for these two paradigmatic chemosensory alterations have been proposed, such as nasal obstruction/congestion and rhinorrhea, damage of olfactory receptor neurons, infiltration of olfactory centers in the central nervous system, and injury of cells supporting the olfactory/taste epithelium [26]. Irrespective of the specific cause, substantial evidence has been provided that self-reported taste and/or smell dysfunctions is associated with community spread of the infection, as well as number of hospital admissions, and are capable of predicting disease epidemiology better than using conventional indicators [27].

The results of our analysis suggest that the volume of Google searches for olfactory and gustatory dysfunctions may help predict the epidemiological trajectory of COVID-19 prior to changes in official reporting. A similar analysis was published by Cherry et al. [28], who reported modest Spearman’s correlations (i.e., between 0.30 and 0.65) between COVID-19 cases and loss of smell or taste. However, these authors limited their search up to May 17, 2020, and only used a seven-day moving-mean. Unlike those findings, we have extended our search up to November 2020, thus also capturing the beginning of the so-called “second wave” of the infection [29]. We have also analyzed the correlations up to four weeks after the index week of Google searches. This has allowed us to define that the volume of Google searches for loss of taste and/or smell in the US efficiently predicted the number of SARS-COV-2 diagnosed cases in the subsequent two weeks.

In conclusion, health organizations and governments should strongly consider using the number of Google searches as an adjunct tool for planning future strategies to prevent, contain, and manage outbreaks of the ongoing COVID-19 pandemic.

## Data Availability

The datasets generated during and/or analysed during the current study are available from the corresponding author on reasonable request.

## Compliance with Ethical Standards

### Human and Animal Rights

No human participants and/or animals and no information consent needed.

### Conflict of Interest

No conflict of interests.

### Ethical Approval

This article does not contain any studies with human participants performed by any of the authors.

### Funding

None

## Acknowledgments

None

## Notes

### Competing Interest Statement

The authors have declared no competing interest.

## References

1. Lippi G, Sanchis-Gomar F, Henry BM (2020) Coronavirus disease 2019 (COVID- 19): the portrait of a perfect storm. Ann Transl Med 8 (7):497. doi:10.21037/atm.2020.03.157

2. Lippi G, Henry BM, Bovo C, Sanchis-Gomar F (2020) Health risks and potential remedies during prolonged lockdowns for coronavirus disease 2019 (COVID-19). Diagnosis (Berl) 7 (2):85–90. doi:10.1515/dx-2020-0041

3. Fontanet A, Cauchemez S (2020) COVID-19 herd immunity: where are we? Nat Rev Immunol 20 (10):583–584. doi:10.1038/s41577-020-00451-5

4. Rubin EJ, Baden LR, Morrissey S (2020) Audio Interview: Guidelines for Covid-19 Vaccine Deployment. N Engl J Med 383 (11):e88. doi:10.1056/NEJMe2029435

5. Lee A, Thornley S, Morris AJ, Sundborn G (2020) Should countries aim for elimination in the covid-19 pandemic? BMJ 370:m3410. doi:10.1136/bmj.m3410

6. Stokes EK, Zambrano LD, Anderson KN, Marder EP, Raz KM, El Burai Felix S, Tie Y, Fullerton KE (2020) Coronavirus Disease 2019 Case Surveillance - United States, January 22-May 30, 2020. MMWR Morb Mortal Wkly Rep 69 (24):759–765. doi:10.15585/mmwr.mm6924e2

7. Lippi G, Mattiuzzi C (2020) Diagnostic and clinical significance of “atypical” symptoms in coronavirus disease 2019. Pol Arch Intern Med 130 (6):478–480. doi:10.20452/pamw.15448

8. Hajikhani B, Calcagno T, Nasiri MJ, Jamshidi P, Dadashi M, Goudarzi M, Eshraghi AA, Facs, Mirsaeidi M (2020) Olfactory and gustatory dysfunction in COVID-19 patients: A meta-analysis study. Physiol Rep 8 (18):e14578. doi:10.14814/phy2.14578

9. Tong JY, Wong A, Zhu D, Fastenberg JH, Tham T (2020) The Prevalence of Olfactory and Gustatory Dysfunction in COVID-19 Patients: A Systematic Review and Meta-analysis. Otolaryngol Head Neck Surg 163 (1):3–11. doi:10.1177/0194599820926473

10. Hoang MP, Kanjanaumporn J, Aeumjaturapat S, Chusakul S, Seresirikachorn K, Snidvongs K (2020) Olfactory and gustatory dysfunctions in COVID-19 patients: A systematic review and meta-analysis. Asian Pac J Allergy Immunol 38 (3):162–169. doi:10.12932/AP-210520-0853

11. Centers for Disease Control and Prevention. CDC COVID Data Tracker. https://covid.cdc.gov/covid-data-tracker/#trends_dailytrendscases. Accessed November 7 2020

12. Cervellin G, Comelli I, Lippi G (2017) Is Google Trends a reliable tool for digital epidemiology? Insights from different clinical settings. J Epidemiol Glob Health 7 (3):185–189. doi:10.1016/j.jegh.2017.06.001

13. Dugas AF, Jalalpour M, Gel Y, Levin S, Torcaso F, Igusa T, Rothman RE (2013) Influenza forecasting with Google Flu Trends. PLoS One 8 (2):e56176. doi:10.1371/journal.pone.0056176

14. Yang S, Santillana M, Kou SC (2015) Accurate estimation of influenza epidemics using Google search data via ARGO. Proc Natl Acad Sci U S A 112 (47):14473–14478. doi:10.1073/pnas.1515373112

15. Crowson MG, Witsell D, Eskander A (2020) Using Google Trends to Predict Pediatric Respiratory Syncytial Virus Encounters at a Major Health Care System. J Med Syst 44 (3):57. doi:10.1007/s10916-020-1526-8

16. Shin SY, Seo DW, An J, Kwak H, Kim SH, Gwack J, Jo MW (2016) High correlation of Middle East respiratory syndrome spread with Google search and Twitter trends in Korea. Scientific reports 6:32920. doi:10.1038/srep32920

17. Mavragani A (2020) Infodemiology and Infoveillance: Scoping Review. J Med Internet Res 22 (4):e16206. doi:10.2196/16206

18. Lippi G, Mattiuzzi C, Cervellin G (2020) Google search volume predicts the emergence of COVID-19 outbreaks. Acta Biomed 91 (3):e2020006. doi:10.23750/abm.v91i3.10030

19. Rovetta A, Bhagavathula AS (2020) Global Infodemiology of COVID-19: Analysis of Google Web Searches and Instagram Hashtags. J Med Internet Res 22 (8):e20673. doi:10.2196/20673

20. Jimenez A, Estevez-Reboredo RM, Santed MA, Ramos V (2020) COVID-19 symptom Google search surges, precede local incidence surges: evidence from Spain. J Med Internet Res. doi:10.2196/23518

21. Peng Y, Li C, Rong Y, Chen X, Chen H (2020) Retrospective analysis of the accuracy of predicting the alert level of COVID-19 in 202 countries using Google Trends and machine learning. J Glob Health 10 (2):020511. doi:10.7189/jogh.10.020511

22. Sulyok M, Ferenci T, Walker M (2020) Google Trends Data and COVID-19 in Europe: correlations and model enhancement are European wide. Transbound Emerg Dis. doi:10.1111/tbed.13887

23. Nakashima T, Suzuki H, Teranishi M (2020) Olfactory and gustatory dysfunction caused by SARS-CoV-2: Comparison with cases of infection with influenza and other viruses. Infect Control Hosp Epidemiol:1-2. doi:10.1017/ice.2020.196

24. Passarelli PC, Lopez MA, Mastandrea Bonaviri GN, Garcia-Godoy F, D’Addona A (2020) Taste and smell as chemosensory dysfunctions in COVID-19 infection. Am J Dent 33 (3):135–137

25. Mullol J, Alobid I, Marino-Sanchez F, Izquierdo-Dominguez A, Marin C, Klimek L, Wang DY, Liu Z (2020) The Loss of Smell and Taste in the COVID-19 Outbreak: a Tale of Many Countries. Curr Allergy Asthma Rep 20 (10):61. doi:10.1007/s11882-020-00961-1

26. Butowt R, von Bartheld CS (2020) Anosmia in COVID-19: Underlying Mechanisms and Assessment of an Olfactory Route to Brain Infection. Neuroscientist:1073858420956905. doi:10.1177/1073858420956905

27. Pierron D, Pereda-Loth V, Mantel M, Moranges M, Bignon E, Alva O, Kabous J, Heiske M, Pacalon J, David R, Dinnella C, Spinelli S, Monteleone E, Farruggia MC, Cooper KW, Sell EA, Thomas-Danguin T, Bakke AJ, Parma V, Hayes JE, Letellier T, Ferdenzi C, Golebiowski J, Bensafi M (2020) Smell and taste changes are early indicators of the COVID-19 pandemic and political decision effectiveness. Nat Commun 11 (1):5152. doi:10.1038/s41467-020-18963-y

28. Cherry G, Rocke J, Chu M, Liu J, Lechner M, Lund VJ, Kumar BN (2020) Loss of smell and taste: a new marker of COVID-19? Tracking reduced sense of smell during the coronavirus pandemic using search trends. Expert Rev Anti Infect Ther:1-6. doi:10.1080/14787210.2020.1792289

29. Grech V, Cuschieri S (2020) COVID-19: A global and continental overview of the second wave and its (relatively) attenuated case fatality ratio. Early Hum Dev:105211. doi:10.1016/j.earlhumdev.2020.105211

